# Approach and Method for Bayesian Network Modelling: A Case Study in Pregnancy Outcomes for England and Wales

**DOI:** 10.1101/2024.01.06.24300925

**Authors:** Scott McLachlan, Bridget J Daley, Sam Saidi, Evangelia Kyrimi, Kudakwashe Dube, Crina Grosan, Martin Neil, Louise Rose, Norman E Fenton

## Abstract

Efforts to fully exploit the rich potential of Bayesian Networks (BNs) have hitherto not seen a practical approach for development of domain-specific models using large-scale public statistics which have the potential to reduce the time required to develop probability tables and train the model. As a result, the duration of projects seeking to develop health BNs tend to be measured in years due to their reliance on obtaining ethics approval and collecting, normalising, and discretising collections of patient EHRs. This work addresses this challenge by investigating a new approach to developing health BNs that combines expert elicitation with knowledge from literature and national health statistics. The approach presented here is evaluated through the development of a BN for pregnancy complications and outcomes using national health statistics for all births in England and Wales during 2021. The result is a BN that when validated using vignettes against other common types of predictive models including multivariable logistic regression and nomograms produces comparable predictions. The BN using our approach and large-scale public statistics was also developed in a project with a duration measured in months rather than years. The unique contributions of this paper are a new efficient approach to BN development and a working BN capable of reasoning over a broad range of pregnancy-related conditions and outcomes.

## 1 INTRODUCTION

Traditional approaches for creating pregnancy prediction models focus on identifying a set of risk factors or symptoms for a singular health issue such as predicting whether the mother may experience gestational diabetes mellitus (GDM) or preeclampsia during her pregnancy. Each model stands alone with no ability to draw on the input data or predictions of other models or the pregnancy as a whole. Such approaches are typical of most predictive modelling in medicine. These models are mostly statistical and often result from a process that seeks those risk factors or symptoms that correlate most with the health issue being modelled. In general, such modelling usually suffers from the following limitations:

- A focus on predicting presence of a condition of interest without considering knowledge of absence of that condition
- Lack of access to sufficient high quality data, resulting in low quality predictions in the presence of uncertain or missing data.
- Use of internal statistical validation that evaluates the model’s prediction but is unable to evaluate the structure or reasoning process.
- Limited case vignettes with exemplar predictions that would help readers relate to the model and compare it to other predictive and diagnostic approaches.

The lower methodological quality and poor reproducibility of many models [1, 2], and authors who simply present the same model in many articles with only minor differences in the cohort [3], have led some to claim that predictive models amount to little more than junk science [4, 5]. While the focus of this study is on the problem of predicting pregnancy outcomes, the proposed approach is sufficiently general that it can be used for creating and validating holistic whole-of-condition predictive models for a wide range of medical conditions.

We propose to address the limitations or typical predictive models by using Bayesian networks (BN). These have already been used for predictive and risk modelling in many domains including healthcare, with comparative evaluations showing BNs produce more reliable results [6, 7]. The approach presented in this paper proposes a complex model that results in a BN capable of reasoning the broad experience of pregnancy for an entire community or nation. This approach draws on: (i) large collections of publicly available statistics; (ii) knowledge elicited from experts; and (iii) published studies. These are used together to construct a novel BN model capable of discriminating the full range from healthy to disease-affected pregnancy, and the probability for different maternal and neonate outcomes. The proposed model is also extensively validated using expert and concurrent analyses.

The paper is structured as follows: Section 1 introduces the motivation, context and research problem addressed in this work; Section 2 presents the theoretical and application domain backgrounds as well as the literature review covering works related to the research problem; Section 3 presents the approach and method developed for the task of knowledge, data and expert-driven modelling of the problem domain space using causal BN; Section 4 presents the results of applying the approach and method to the case for developing the BN for the medical domain of pregnancy complications and outcomes; Section 5 discusses the experience of using the approach and method, and identifies potential limitations that may arise when applied to other problem domain spaces.

## 2 BACKGROUND AND LITERATURE REVIEW

We screened a collection of 100 works published between 2000 and 2023 that proposed a predictive risk screening model for pregnancy complications, including preeclampsia (PE), gestational diabetes mellitus (GDM), preterm birth or stillbirth [8].

### 2.1 Risk Factors and Symptoms

Clinical risk screening for adverse pregnancy outcomes commonly occurs during the first maternal clinic appointment, which is often described as the booking visit [9–11]. While the risk screening score may be updated during antenatal care as the pregnancy progresses and new clinical and non-clinical information arises [12], a variety of different signs, symptoms or clinical tests may be involved dependant on whichever clinical guideline or scoring model has been adopted. When identifying the range of clinical and risk factors that best fit their model’s aims, some rely on different combinations of common factors like maternal age, BMI and pregnancy history collected during the booking visit [9, 11], while others retrospectively draw on a broad range of antenatal care records and the pregnancy outcomes [13, 14]. Some authors include what are commonly called booking bloods (a range of normal blood tests ordered prior to or during the booking visit that includes capillary blood glucose (CBG), fasting plasma glucose (FPG), or a full blood count (FBC) [15] while others incorporate antenatal screening tests such as ultrasounds or oral glucose tolerance tests [15, 16]. Others even introduce novel or uncommon variables such as paternal DNA [17] or a vaginal swab for bacterial vaginosis [18] in order to identify whether their addition improves predictive accuracy. A list of the different risk factors and symptoms used in their predictive models by authors in the collection we screened is provided in Table 2 of Appendix 1.

### 2.2 Common Issues

Several issues have frustrated many seeking to create healthcare risk, probability and decision support models.

First, collecting a dataset of sufficient size and quality healthcare records capable of supporting complex and broad understanding of the incidence, risk factors, causes, treatments and prognosis of a disease remains an intractable issue [19].

In our literature review we saw many models for diagnosis or prognostication based on the records of very small patient groups. While the data for a model may set out with several hundred or even thousands of patients, when factored based on particular demographic or clinical risk factors, there may be as few as 1, 7, or 9 [20], 14 [12] or 23 patients [19] remaining in key diagnostic subgroups for ingestion to train the model.

Second, evaluation and validation of predictive models has traditionally been limited to internal statistical approaches that distinguish between positive and negative cases: archetypal statistical and graphical methods such as: (i) the receiver operator curve (ROC); (ii) p-values; and (iii) confidence intervals (CI). The ROC curve plots the true positive rate (TPR) (sensitivity) against the false positive rate (FPR) (1-specificity or the probability of a false alarm) at various threshold levels. Many results are also presented with either p-values, CI, or both, which the authors submit is a measure of the strength, or statistical significance, and degree of confidence we might have in their results. These statistical approaches essentially seek to compare model predictions to data available for the subject matter, which in some cases is also the data the model was trained with. Our screening review found that most prediction models for pregnancy-related health issues were statistical, rather than AI, and that half of the reviewed works also used these three common statistical approaches [8]. It has been argued that the ability of common statistical methods to accurately test the validity of a BN is limited, and that more rigorous frameworks should be employed for this purpose [21].

A key difference between the models we reviewed, and the model presented in this work, is that the reviewed models focus on identifying the most predictive collection of symptoms and risk factors for an individual outcome from a dataset of retrospective patient records - for example: which symptoms and risk factors are most predictive for pre-eclampsia? In contrast, the model presented in this work is a causal model – a BN - built using extensive statistics and expert elicitation, and combines many such individual outcomes in order to provide an overall representation of pregnancy and pregnancy outcomes.

Third, is that accuracy of predictions of most models can vary wildly when: (i) observations of some risk factors, signs or symptoms are missing; or (ii) when the patient does not actually have the health condition that the model is designed to predict. In the first instance the model may underestimate risk for the patient having the condition, while in the second it may overestimate. These issues are caused by overfitting that many machine learning (ML) and algorithm-based solutions inevitably suffer from. These solutions are developed with knowledge or trained with data that identifies what is the medical condition under investigation, often to the complete exclusion of anything that would inform the model as to what isn’t that medical condition. Over-fitting is seen where authors exclude participants who either do not have the disease of interest, or who were found to have other similar diseases. When presented with a broader dataset, these solutions tend to identify the disease of interest in patients who actually have other conditions that may share even only a small number of similar symptoms, or present in academic papers with claims of unreasonably high degrees of accuracy from a very limited number of data points when tested with the author’s training dataset. For example, in development of an automated ML model to identify gestational diabetes mellitus (GDM) [22] excluded women of black, white or mixed ethnicities, those who were found to have the clinically similar though not identical Type 2 Diabetes Mellitus (T2DM) in pregnancy, and those whose glucose tests were reported outside a narrow 4-week window from their dataset; and based on analysis of only four data points (HbA1c, mean arterial blood pressure, fasting insulin and the woman’s HDL ratio) and testing with their own training dataset of 222 participants, the authors claimed an ‘excellent’ 93% accuracy for preconception prediction of future GDM.

There was a final issue observed of most works proposing a predictive model. Some works focused, sometimes with excessive detail and to the almost complete exclusion of anything that would allow the reader to easily judge the model’s performance in a real-world clinical situation. Their focus was either on describing: (a) the incidence of individual risk factors that feed into their model; (b) the complex algorithm that forms the basis of their model; or (c) the AUC/ROC or accuracy results of their chosen approach. Many works in the literature presented some or all these three absent an easily understandable example, such as a vignette of a risk scored case or probability scores generated through operation of their model. We believe inclusion of such exemplar cases would help make the proposed model more approachable and comprehendible to clinical readers, potentially increasing adoption of predictive and clinical decision-support models in clinical practice. We only identified two works that provided such a vignette and risk score. First, [20] developed a risk prediction mode for preeclampsia (PE) in first-time mothers that includes a vignette and risk prediction (%) for diagnosis of PE for a 28-year-old woman with high mean arterial blood pressure (MAP) and body mass index (BMI) and a family history of PE. Second, [23] describes an approach for predicting premature birth using a range of commonly recorded risk factors and include a short vignette example and risk prediction (% for preterm delivery for a 35-year-old first-time black mother who smokes but has no prior diagnosis of diabetes. These examples help to ground the author’s model and make it approachable for readers who are not expert mathematicians or biostatisticians. They also provide an easily identifiable comparator that can be the starting point for evaluation or validation against other models.

## 3 METHOD

Just like many of the papers we reviewed, when it came to developing causal models to support risk estimation and clinical decision-making for conditions like rheumatoid arthritis, angina, acute traumatic coagulopathy and GDM our research group initially focused on individual diagnoses, outcomes or clinical decisions [24–26]. We also initially failed to consider either construction of a model representing general health when the disease of interest was not present, or of the scope of observable experience for a population within a health service, district or entire country. We now believe that it is only once AI developers can model a more holistic view of the patient, their community, and disease, that we can truly begin to develop and demonstrate the credibility of our models: (a) identifying or explaining risk; or (b) providing computer-based (ML or AI) clinical decision-support. Further, we believe that it is only once we have established a community-wide baseline that we can evaluate causal relationships between both previously identified and novel symptoms (is this new symptom impacting on or caused by the disease of interest?), and model treatment and prognostic counterfactuals (what would happen if we administered this treatment for this patient?).

### 3.1 Hypothesis

The hypothesis for this work is that it is possible to develop a broadly accurate BN model capable of describing diagnosis and treatment outcomes using expert clinical knowledge, large publicly available privacy-preserving datasets, and published statistics for a given population. Commonly recorded observations made about the medical condition of interest could be made on this model to derive predictions regarding health condition outcomes. This model would incorporate clinical knowledge about causal interactions between clinical knowledge, patient information and publicly available clinical data and could also be used provide prospective risk and outcome predictions. To the best of our knowledge a BN modelling approach that uses large-scale health and outcome statistics has not previously been undertaken.

### 3.2 Bayesian Networks

Bayesian networks (BNs), also known as *probabilistic graphical models* or *belief networks*, are a directed acyclic graph (DAG) that provide a graphical representational framework for probabilistic reasoning under uncertainty. The BN comprises two main *components, structure* and *parameters*. The structure is made up of variables, or nodes, representing random variables (discrete or continuous) and directed edges representing causal or influential relationships. If a directed edge connects variables A and B, such as A → B, then A is called parent node or ancestor of B, and B is a child node or a successor of A. The BN parameters comprise a set of conditional probability functions associated with each node to represent the strength of each node in the BN given its parents. Bayesian probabilistic reasoning describes the process of updating our prior belief about an uncertain hypothesis by considering new evidence. Our initial belief is termed prior probability, while our updated belief is termed posterior probability.

The process by which the probabilities of the entire BN are updated is often described as reasoning or inference. Figure 1 presents several scenarios wherein observations are made and outcomes observed on a simple model for lung cancer. For each example in Figure 1 the observed evidence entered into the model is shown in grey, while the nodes whose updated probability determines the question being reasoned about are shown in black. The top two examples in Figure 1 demonstrate reasoning from evidence in two directions:

- **Forward reasoning** follows the direction of the arc. Being a smoker is a predisposing factor that influences the likelihood for whether an individual will be diagnosed with lung cancer and have a positive diagnostic X-ray and dyspnoea. Forward reasoning is described as *causal* or *predictive reasoning* when the BN structure represents true causal relationships rather than simple associations, which is not an absolute requirement when we reason from evidence.
- **Backward reasoning** operates counter to the direction of the arc. If we identify that the patient has dyspnoea, this observation will update our prior belief that the patient might have cancer and may have been previously exposed to high levels of pollution. Background reasoning is also described as *diagnostic reasoning* when the BN structure represents true causal relationships.

**Figure 1:**
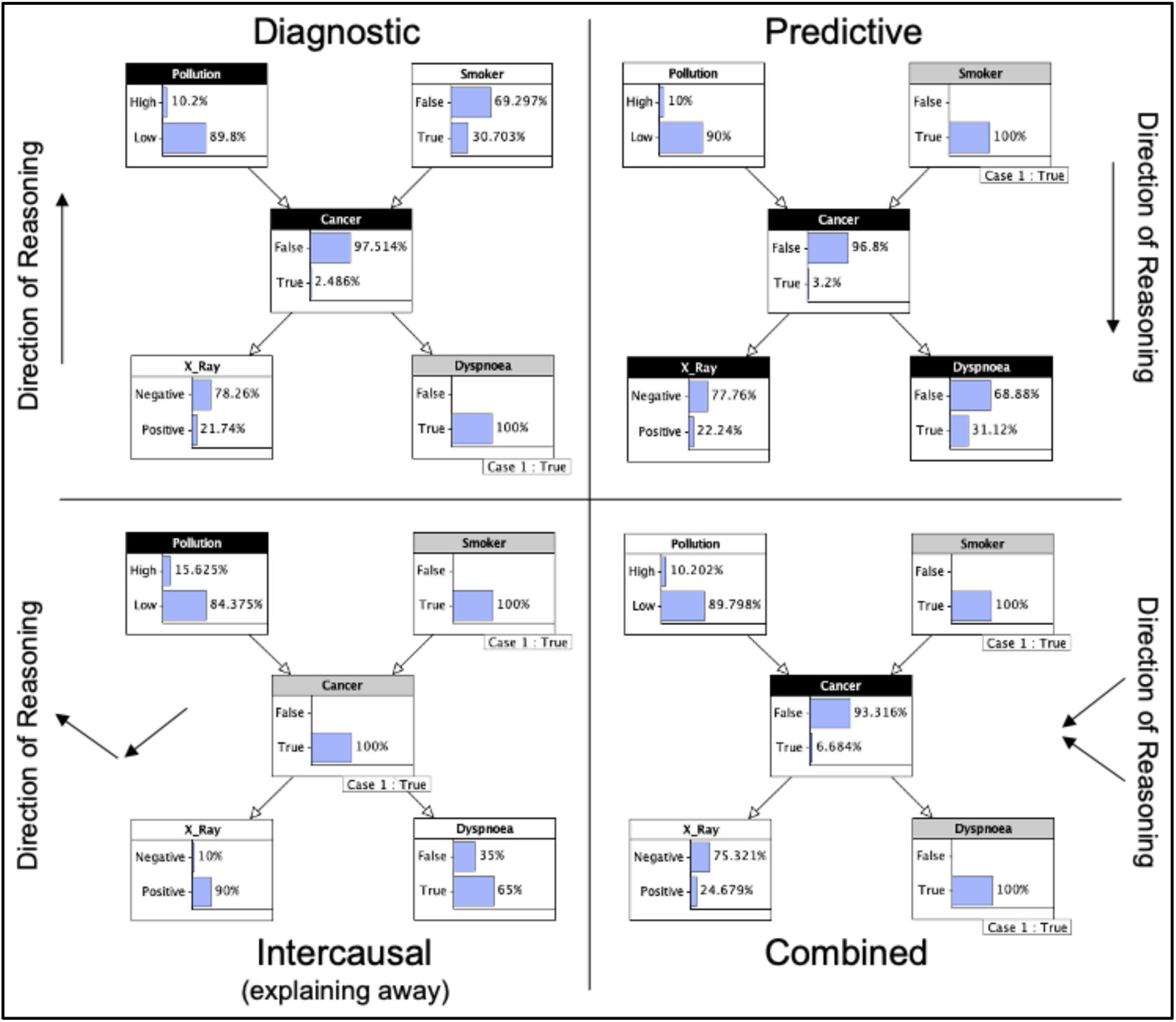
Types of reasoning

The bottom half of Figure 1 also shows that it is possible to combine forward and backward reasoning in the same reasoning process:

- **Intercausal reasoning** describes a cross-reasoning situation where the forward reasoning observation of one parent node along with observation of that parent’s child, through the application of backward reasoning, explains away the unobserved parent nodes of the same child. In this example we have observed one risk factor (smoking) and know the patient has the disease (cancer), which together reduce the significance of or explains away the potential of other unknown risk factors (pollution).
- **Combined reasoning** combines both forward, intercausal and backward reasoning toward a common output. In this example forward (predictive) and backward (diagnostic) reasoning are combined to ascertain the probability for whether a patient with both a symptom (dyspnoea) and a risk factor (smoking) has the disease (cancer).

In total, four different modes of reasoning are possible from this type of BN. Our aim is that the model presented in this work should also be capable of performing all four modes of reasoning.

### 3.3 Study Population

The data for the model presented in this work were derived from the following publicly available privacy-preserving statistical sources:

*The United Kingdom (UK) Office for National Statistics (ONS):*

- Birth Characteristics 2021 dataset [27]
- Child and Infant Mortality 2021 dataset
- Preconception health among migrant women in England 2019-2021 dataset

*Mothers and Babies:* Reducing Risk through Audits and Confidential Enquiries across the UK (MBRRACE-UK)

- MBRRACE-UK Perinatal Mortality Surveillance: UK Perinatal Deaths for births from January to December 2021

*Public Health England (PHE) National Congenital Abnormality and Rare Disease Registration Service (NCARDRS)*

- NCARDRS Congenital abnormality statistics: Annual data 2021 dataset [51]

These sources describe a broad range of risk factors and their incidence for the following events that occurred in England and Wales during 2021:

- 624,828 pregnancies (including 8,470 multiparous pregnancies)
- 624,162 live births
- 2,579 stillbirths
- 1,353 neonatal deaths under 7 days
- 1,715 neonatal deaths under 28 days
- 608 post-neonatal deaths 28 days or over
- 312 post-neonatal deaths between 4 weeks and 3 months
- 160 post-neonatal deaths between 3 and 6 months, and
- 136 post-neonatal deaths between 6 months and 1 year

Additional evidence for establishing incidence and causal relationships between risk factors and symptoms were derived from clinical practice guidelines [28] and academic studies of pregnancy and pregnancy outcomes published between 2019 and 2022. Emphasis was given to those studies that described data collected in 2021 from UK populations.

### 3.4 BN Development

Our primary design goal is that the model should represent a credible overview of current clinical knowledge regarding key risk factors and signs and symptoms that interact to impact upon pregnancy outcomes. To achieve this goal a six-phase development process was adopted and is as illustrated in Figure 2. The rest of this section explains these phases.

**Figure 2.**
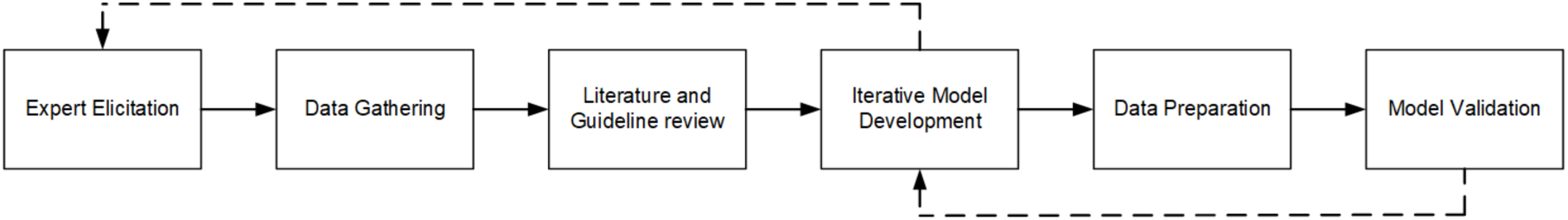
BN Development Process Flow Diagram

#### 3.4.1 Expert elicitation

It is possible to conceive the structure and parameters of the BN model entirely from data in situations where an extensive dataset with contextual information is available. However, BNs provide a flexible platform that can easily combine less comprehensive datasets with multiple expert’s knowledge and other disparate sources of information [29]. Expert input benefits the design process and enhances the resulting BN by ensuring the complexities of up-to-date domain knowledge can be incorporated [30, 31]. The expert elicitation process used in our work created information visualisations called caremaps that describe the progression of disease, diagnosis and treatment processes and possible patient outcomes in the form of a flow diagram or process map. The process for developing caremaps containing clinical decision points and decision criteria has previously been described in [32]. Our clinical experts for this work included a registered midwife with more than 15 years experience (BJD) and an associate professor and specialist in obstetrics and gynaecology (SS).

#### 3.4.2 Data gathering

We sought national datasets on pregnancy that described the incidence of pregnancy complications and outcomes for an entire population. A key focus for this work was to seek publicly available privacy-preserving datasets whose use would not require or violate institutional ethics policies. This limited us to secondary or aggregate statistical sources such as those of national health services, government health departments or national statistics agencies. We collected datasets for the year 2021 because these were the most recent complete statistical datasets for the UK available from government and academic sources at the time.

#### 3.4.3 Literature and Clinical Guideline Review

We conducted a literature search to locate literature, clinical practice guidelines, and protocols relevant to the medical condition(s) being modelled. The literature included is then aligned to the data that has been gathered in Phase 3.4.2 above. Priority is given to those articles published during the period matching the data collected in the previous phase, which describes incidence of the medical condition(s) recorded from like populations.

#### 3.4.4 Model development

An iterative model development process was identified wherein: (1) medical idioms that identify key structural fragments necessary to the model would be identified from a combination of the caremap and knowledge derived from the clinical experts; (2) data would be identified from the statistical and academic sources that first described incidence of the variable of the node and second the incidence of interaction across the arcs between that node and its parent or child nodes to populate node probability tables (NPTs); (3) the structural fragments would be brought together into a single contiguous BN model; and (4) the resulting model structure would be reviewed with the clinical experts and where changes are identified, the process would return to the first step. The process for identifying medical idioms and using idioms to expedite expert elicitation-supported construction of BNs has been previously described in [33].

#### 3.4.5 Data preparation

Each node within a BN has a NPT. Absolute parent nodes (parent nodes that are not also children of another node – such as the pollution node in the example in Figure 1) have a single dimensional NPT. Where a node has a single parent (such as the dyspnoea node in the example in Figure 1) it will have a two-dimensional NPT. Where a node has two parents (such as the cancer node in the example in Figure 1) it will have a three-dimensional NPT. This continues up to nodes with six parents. Nodes with six parents or greater are generally avoided not only because it is possible to incur considerable computational penalty during model operation, but primarily due to the elicitation burden they impose on experts and the potential impossibility of performing the complicated mental math required to resolve NPT at such high dimensionalities [34]. Examples of data dimensionality in NPT are provided in Figure 3.

**Figure 3:**
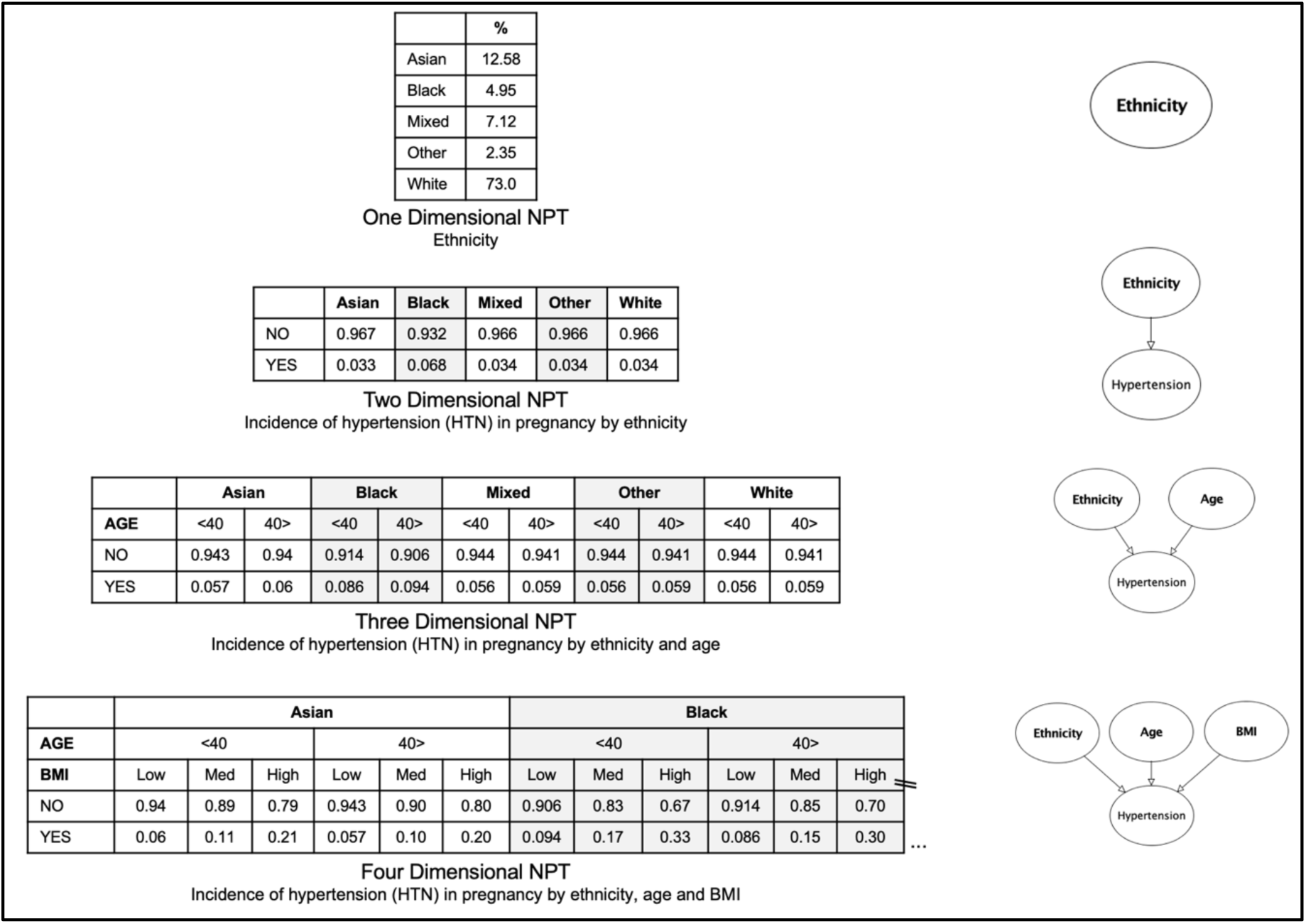
Examples of dimensionality in NPT

The content of each column in an NPT should sum to 1.0 (100%). Discretisation allows the modeller to convert continuous variable factors like BMI and capillary blood glucose (CBG) by assigning them *clinically relevant intervals,* ordinal states or categories (for example: low, medium, high). Some variables such as BMI in the example shown in Figure 3 were discretised. Tables using population-level continuous variable data were prepared in Microsoft Excel and converted on ingestion by the Agenarisk BN modelling tool.

#### 3.4.6 Model validation

We chose a more extensive multi-step validation process recommended by several authors [21, 35, 36] for validating BN models. We commenced by using an approach that allows for the model structure and degree to which interconnected variables interact to be iteratively validated during construction with clinical experts (face validity). Face validity is one of the most common validation tests for graphical models like expert-elicited BNs, but ironically it can also be the weakest [21]. The key weakness arises when you seek to establish face validity from experts who were involved in the model’s design, because they are unlikely to disagree with a model that reflects their own judgement [21]. While we acknowledge that there may be more risk factors and relationships than it is possible to model, or that new risk factors may be identified by future research, we will use our screening review and relevant clinical practice guidelines to ensure the most important or significant risk factors and symptoms are included (content validity). Content validity tests whether the BN structure contains key factors, how they are represented, and that relationships identified from the literature have been included in the model [21]. Content validity also seeks to identify those relationships that are new and therefore only found in the proposed model. We will also seek to identify clinical vignettes that describe reasoning examples using models in the literature that predict similar outcomes, testing the predictions of those models against the related fragment within our model (concurrent validity). Concurrent validity tests whether the BN, or a boundable fragment of the BN, performs comparatively to another approach seeking the same outcome [21].

## 4 RESULTS

The BN model for pregnancy outcomes is shown in Figure 4. The model is divided into four fragments:

- **Maternal:** This fragment contains the mother’s demographics, pregnancy-relevant risk factors and medical history that are collected during her initial midwifery clinic appointment (in army green), relevant blood tests performed during that first clinic appointment (in red), and nodes representing three health conditions that are common and therefore specifically monitored for in pregnancy (in purple).
- **Birth outcome:** This fragment contains factors and outcomes relevant to birth and initial survival of the baby.
- **Maternal outcome:** This fragment contains several patient and family history factors collected during the initial clinic appointment (in army green), postnatal symptoms (in purple) and health conditions (in white) that have potential to negatively impact the mother’s survival.
- **Neonate outcome:** This fragment considers family history (in army green), infection (in light blue), and health issues that sometimes are only identified (in dark green) or occur (in pink) postnatally and act upon survivability for live-born babies.

**Figure 4:**
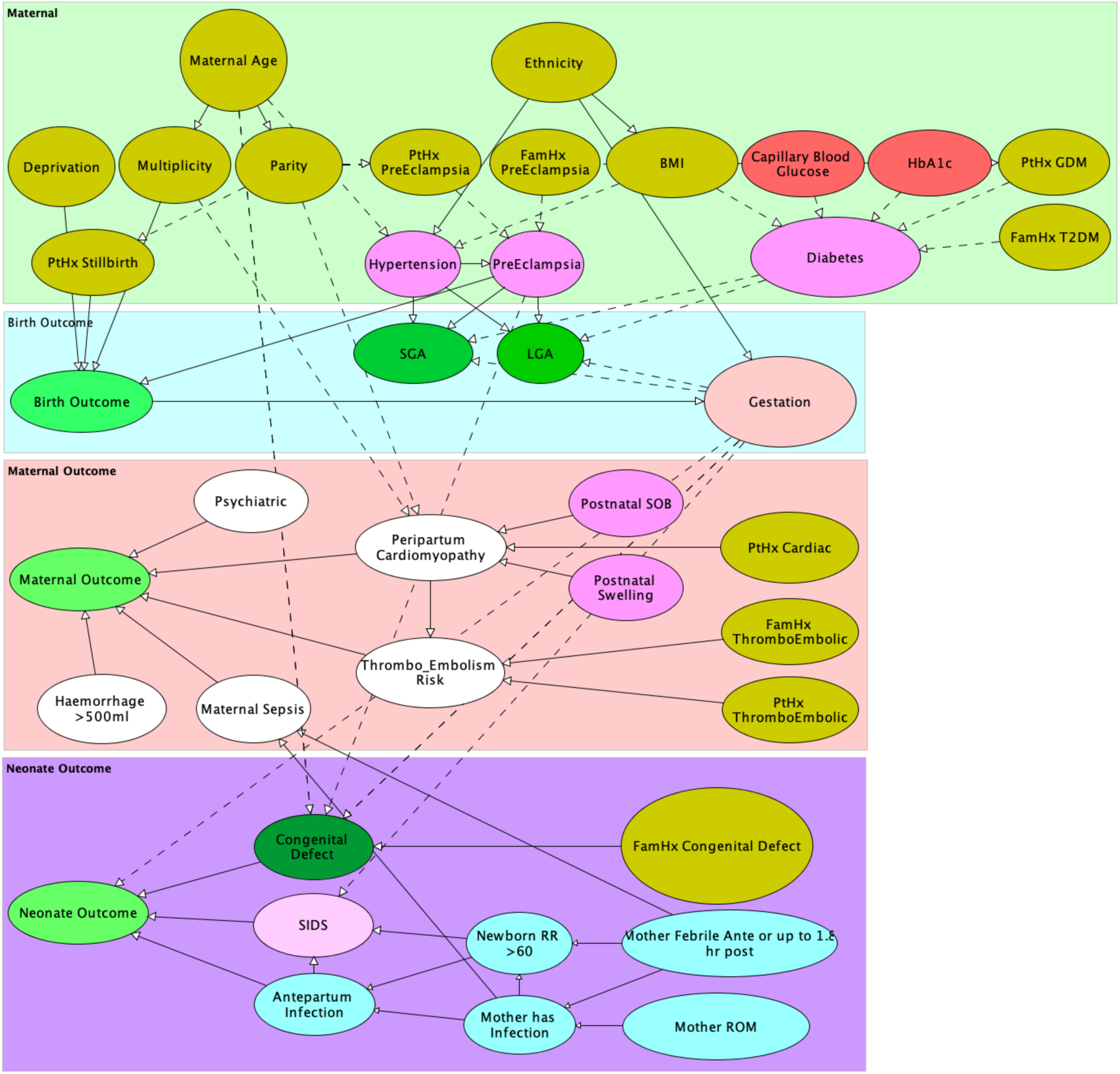
BN model for pregnancy outcomes

The model structure allows the impact of evidence observed to nodes in one zone to affect child nodes in other zones. This ensures the impact of aspects observed during the pregnancy (in the Maternal zone) or birth (in the birth outcome zone) that reduce potential survivability for the neonate to propagate and update the baseline priors of the maternal outcome and neonate outcome zones. The arcs between nodes are variously shown as solid or dashed lines. Solid lines represent a direct relationship in the direction of the arrow between two nodes. Dashed lines indicate that a hidden node exists along the line between the parent and child node. As will be shown in section 4.1.2, these hidden nodes were used to provide an alternate discretisation of the variable and values described in the parent node. The model priors are shown in Figure 5.

**Figure 5:**
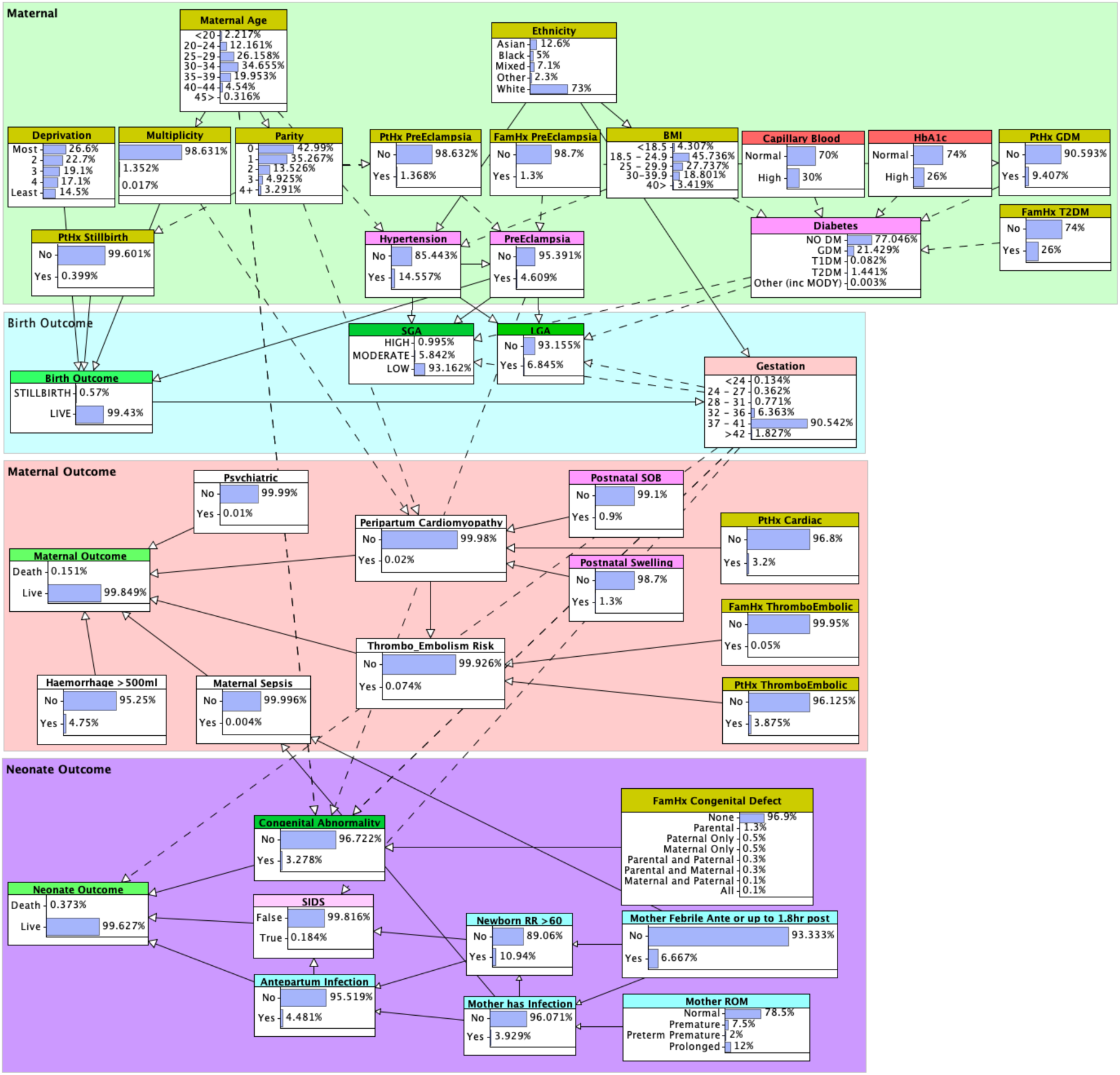
BN model showing background priors

### 4.1 BN Validation

This section provides an overview of the processes used to validate the pregnancy risks and outcomes model.

#### 4.1.1 Face validity

We drew on a small group of experts during the design of structural components of the model, and when those experts agreed with the resulting structure and interaction between linked nodes it was evaluated by comparison to literature and clinical guidelines and by other experts who had not been involved in the design process for that structural component. Face validity asks: does the model look and appear to behave like we expect it to?

- **Structure:** Our model was iteratively developed with ongoing input from clinical experts. The iterative development process sought to ensure that variables and causal pathways arising out of the physiological processes taking place in the mother and baby were weighed against the evidence from clinical texts and medical journals and the data that was available. The goal was to ensure that whenever possible and practicable, and wherever data or statistics was found to support the mechanism of influence, the relationships between physiological processes were represented appropriately in the model. In this way face validity of the structure was evaluated and changes made where necessary at several key stages during the development process.
- **Parameterisation:** Clinicians were asked to provide estimates for BN parameters; both as a background (prior) value as well as to quantify the impact of other observations on the model. For example, we might ask: What is the probability of any woman in the population being diagnosed as GDM? or What is the probability of an underweight Asian woman or overweight Black woman being diagnosed as GDM during her pregnancy? In cases where the model’s parameters that had been learned from national statistics or recent studies were not in agreement with the experts estimates, further investigation was performed. Midwives’ estimates were more often closest to the parameter values observed in national statistics and the literature. Overall, greater weight was given to the national statistics because they reflect what actually happened for the entire population.

Three primary pregnancy outcomes were identified and are represented by the nodes in bright green in the final model:

- **Birth Outcome:** There are two categories for birth outcome - live birth or stillbirth. The reported percentage of stillbirth in the ONS Birth Characteristics 2021 dataset is 0.42%. The prior probability for stillbirth in our model is a slightly higher 0.57% because we elected to include late term miscarriage in the Birth Outcome node. We did this because while most texts define pregnancy loss before 20 weeks gestation as miscarriage, and pregnancy loss after 20 weeks gestation as stillbirth [37, 38], we identified that some authors and NHS facilities count pregnancy loss up to gestation week 24 as miscarriage [39, 40].
- **Maternal Outcome:** This represents the likelihood for maternal death. In 2021, 94 births resulted in maternal death which are correctly reflected in the priors of this node (94/624,162 or 0.0151%).
- **Neonate Outcome:** In 2021, 2,323 (1,715 neonates and 608 post-neonates) deaths occurred in live-born babies (2,323/624,162 or 0.37%).

Three secondary outcomes related to the baby were also identified and are represented by the nodes in dark green in the final model:

- **Small for Gestational Age (SGA):** Whether the child is of naturally small size or the impact of conditions like maternal hypertension that cause foetal growth restriction (FGR) and thus increase infant morbidity and mortality. In the UK an SGA baby is one whose birthweight is below the 10th percentile on a customised growth chart [41, 42]. Our SGA node uses three categories: LOW for babies who are at low risk of being SGA; MODERATE for babies who are at moderate risk of being SGA at or below the 10th centile; and HIGH for babies at high risk of being SGA and at or below the 2nd centile. Reported incidence of SGA in England in 2021 was 6.8% (of which 0.9% were at or below the 2^nd^ centile), while incidence in Wales was 6.0% (of which 1.0% were at or below the 2nd centile). In both cases the incidence figures had been rounded to one decimal place by the authors of the 2021 National Maternity and Perinatal Audit [43]. Our model learned from data for both England and Wales returned priors of 5.84% for MODERATE and 0.99% for HIGH, summing to a total incidence for 2021 of 6.83%.
- **Large for Gestational Age (LGA):** Women who are diabetic or have a high BMI without hypertensive disorders may have an LGA (macrosomic) baby. Babies are classified as LGA when their birthweight is over the 90th percentile on a customised growth chart. Being LGA increases the risk of birth complications both for the baby (e.g.: shoulder dystocia) and for the mother (e.g.: perineal tearing). While incidence of LGA babies reported in studies can range between 2.7% [44] and 20% [45] of all live births, we found NHS published rates from 5% [46] to 13% [47]. Our model learned from 2021 datasets shows a prior of 6.8% which rises to 13.2% if the mother was observed to have gestational diabetes mellitus (GDM).
- **Congenital Abnormality:** Congenital defects are observed as a wide range of genetic (e.g.: Downs, Turners and Marfan’s Syndromes) and non-genetic disorders (e.g.: transposition of the great arteries and medication-associated defects such as thalidomide teratogenesis) that can affect the developing foetus. The likelihood for congenital abnormality increases where a family member has also had a diagnosis [48, 49], and can even be affected by the age of the parents [50]. While the most recent dataset describes a total incidence of 2.22% for both genetic and non-genetic congenital abnormalities identified at birth [51], other UK research has found that when diagnosis in infancy is included rates in some populations in England could be as high as 4.1 - 5.3% [52, 53]. Learned from UK national statistical data and incorporated research regarding increasing prevalence rates due to inheritance within family lines, the prior probability in our model resolved to 3.278%.

#### 4.1.2 Content validity

The most common demographic risk factors identified in works proposing predictive models were: (i) maternal age (74%); (ii) BMI (59%); (iii) parity (42%); (iv) ethnicity (36%); and (v) gestation (32%).

**Maternal Age:** Some works only considered maternal age as a risk factor if the mother was either above or below a particular boundary age [54, 55]. Others incorporate age into their models as a continuous variable [39, 56]. Yet more discretised age into intervals, most commonly spanning five or ten years [57, 58]. Our model discretises age into five-year increments. These increments were sufficient to the required degree of model accuracy and consistent with the national maternity statistics [27] and studies that described the impact of maternal age on pregnancy outcomes related to factors such as parity and multiplicity.

In some cases, it was necessary to simplify the BN structure and reduce NPT complexity in child nodes by including hidden nodes that provide an alternate discretisation of the states of a variable. For example, to simplify representation of the effect of advanced maternal age to significantly increase incidence of conditions such as gestational hypertension [59, 60] we included a hidden boundary age node (<40, 40>) as shown in Figure 6.

**Figure 6:**
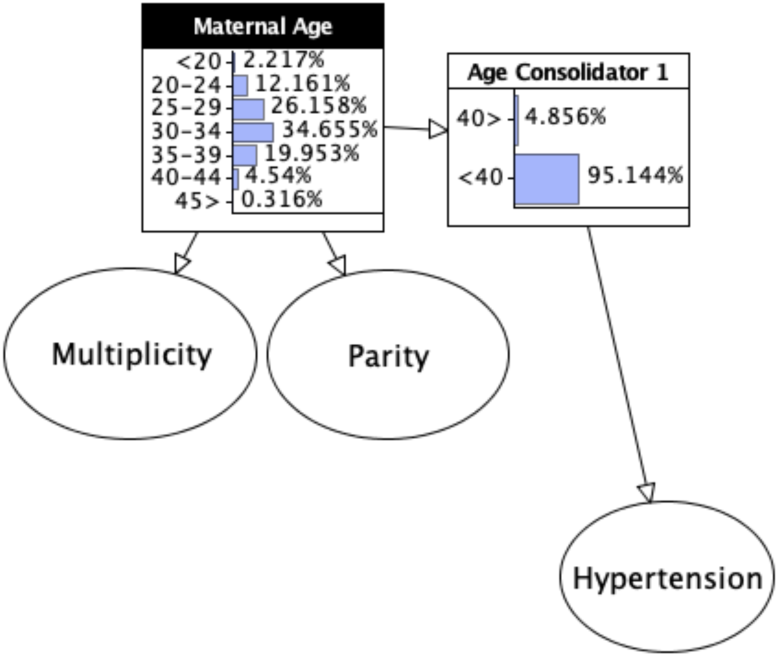
Maternal Age substructure showing hidden Age Consolidator node

**BMI:** The most useable population-wide BMI data relating to pregnancies in England and Wales was found in Public Health England’s *Health of Women before and during pregnancy antenatal booking report* [61]. This report provided BMI statistics in five ranked categories (<18.5, 18.5-24.9, 25-29.9, 30-39.9, 40>) and classified by maternal age, deprivation, and ethnicity. It has been long established that: (i) BMI is a risk factor for GDM [62]; (ii) a relationship exists between ethnicity and maternal weight (represented as BMI) and the incidence of gestational diabetes [63, 64]; and (iii) maternal BMI is an independent risk factor for gestational hypertension that increases the chance of preterm delivery [65, 66]. Elevated maternal weight increases the risk of preterm birth and other maternal complications [65]. Along the obesity-prematurity pathway are GDM and gestational hypertensive disorders [65, 67]. For these reasons we situated BMI between the parent node ethnicity and the child nodes diabetes and hypertension. From this position the impact of BMI can also be seen to flow onto the outcome nodes that describe the likelihood for preterm birth (gestation of less than 36 weeks) and delivery of an SGA or LGA baby [66].

**Parity:** At each extreme of the scale, nulliparity (no previous live births) and grand parity (variously defined in the literature as being the fifth, sixth or seventh pregnancy that results in a live birth) can both be risk factors for pregnancy disorders and outcomes [68]. Parity was provided in UK national datasets [27] in five ranked categories (0, 1, 2, 3,4+). Maternal age is a linked factor, especially where the nulliparous pregnant mother is extremely young or advanced in age [69, 70]. The prior probabilities of nodes such as diabetes, hypertension and birth outcome also update consistent with incidence rates observed in the 2021 dataset [27] when nulliparity or grand parity are observed; including that the model is consistent with the incidence of GDM in mothers with no other risk factors in the 2021 dataset during a first pregnancy (19.6%) versus grand parity (22.5%). Observation of nulliparity on the model is also seen to correctly withdraw any potential impact from other patient history nodes relating to conditions of a previous pregnancy (PtHx GDM, PtHx Stillbirth and PtHx Preeclampsia).

**Ethnicity:** Ethnicity has been identified as a factor that may alter the likelihood and severity of a number of different pregnancy-related conditions and outcomes. These changes may be related to genetic differences, or attributable to different environmental factors such as where the mother lives, her deprivation status, or cultural dietary factors. Ethnicity is a risk factor for gestational and chronic hypertension [71]. The interaction between ethnicity and advanced maternal BMI results in higher rates of GDM and caesarean section for obese Asian mothers, and higher rates of gestational hypertension and caesarean delivery and lower rates of LGA for African mothers [72]. Further, research suggests ethnicity and the gestational age of the baby at birth are risk factors affecting incidence of several poor birth outcomes including chorioamnionitis, birth defects and infant mortality [73, 74]. For these reasons we have focused on the influence of ethnicity on hypertension, gestation and BMI, and through those connections ethnicity’s influence impacts every potential pregnancy outcome computed by the model.

**Gestation:** When the baby is delivered is both an outcome, because it means the pregnancy and birth phases are over, and a risk factor. For example, prematurity, and whether the neonate is at risk of the wide variety of health issues that come from being premature, is decided on the basis of a gestational age at or below 37 weeks at birth. Studies suggest prematurity is a risk factor for developmental, behavioural, emotional, chronic cardiovascular and congenital conditions and infections that can see the neonate requiring extended hospital stays during their first years of life [75, 76]. At the other end of the scale there is increased risk to both mother and baby from being induced or requiring a surgical delivery (caesarean section) for a post-dates (after 42 weeks gestation) pregnancy [77–79]. Gestation is seen to alter the likelihood of a child being SGA or LGA, and the risk that they will either be stillborn or die while they are still a neonate (within the first 28 days after birth). In order to ensure the impact of gestation on maternal and neonate risks and outcomes our gestation node was learned from the 624,162 births in the 2021 dataset [27] and is connected with other birth outcome nodes including SGA, LGA, congenital abnormality, SIDS risk and neonate outcome.

#### 4.1.3 Concurrent validity

We re-examined the papers included in our screening review [8] to locate any works that included a vignette with prediction suitable for use in concurrent validity testing, identifying only two.

[20] propose a model using statistical methods to predict probability of incidence for pre-eclampsia. Their model is based on demographic and risk factors of the first-time mother, along with observable signs and symptoms routinely collected by the midwife during the initial (booking) patient appointment. They used their model to compute a probability for the following vignette:

> *A 28 year old nulliparous woman whose birth weight was 2400 g, with a mean arterial pressure of 96 mmHg, BMI 30, a family history of pre-eclampsia, and no protective factors, her probability of pre-eclampsia is 39%.*

When we observe those elements from the vignette that are also present within the model (maternal age, BMI, parity, family history of pre-eclampsia and maternal hypertension), Figure 7 shows that our model computes a 43.25% probability of pre-eclampsia. We attribute the 4.25% difference to several factors: (i) their dataset included births from multiple disparate countries with different indigenous populations (England and Wales, Ireland, New Zealand and Australia) while our model was built using the statistical data of births that occurred just in England and Wales; (ii) their data reports pregnancies delivered between 2004 - 2008 while the pregnancies in the dataset used to construct our model all delivered in 2021; (iii) their dataset includes 4961 pregnancies while ours includes a significantly larger 624,828; and (iv) many countries report increasing incidence of several pregnancy-related health conditions including gestational hypertension and pre-eclampsia between 2010 and 2020 (Cameron et al, 2022; Sutan et al, 2022).

**Figure 7:**
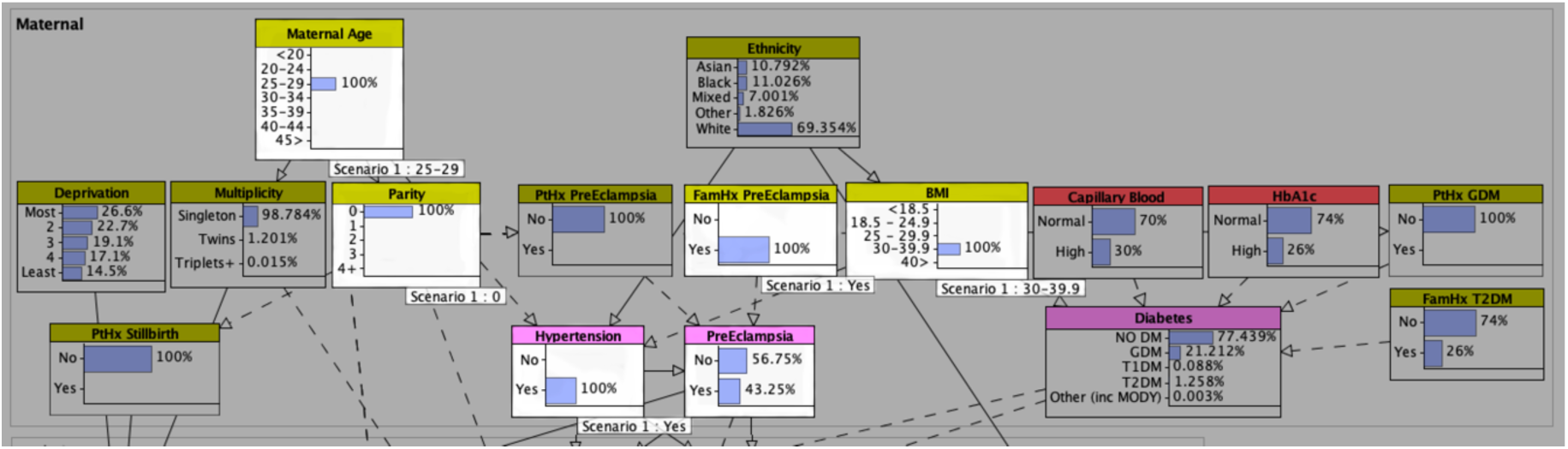
Observation of elements of the vignette from [20]

[23] used statistical methods on pregnancy data collected between 2004 - 2009 to develop a nomogram for predicting preterm delivery. Their dataset began with a larger number of potential factors, while the resulting nomogram includes only the nine factors they resolved as the most predictive from their cohort of 192,208 pregnancies from the United States of America (USA). Their vignette describes:

> *A 35 year old (13 points) African American (41 points) woman planning to get pregnant for the first time (46 points) who has no history of diabetes (0 points) but who smokes (12 points) would have a total of 112 points. This approximates to a baseline probability of preterm birth of 12-13% prior to conception.*

Our model based solely on births in 2021 in England and Wales returned an 8.715% preterm birth prediction – which is the sum of the predictions of all gestations prior to 37 weeks as shown in Figure 8. Smoking was one risk factor absent from our model that was included in theirs that could potentially explain the difference in prediction. Smoking has a significant social stigma attached to it today which has been seen to decrease patients’ willingness to accurately self-report [80, 81]. This can significantly bias the impact of smoking in studies [81]. Smoking rates in different countries vary dramatically, and studies have demonstrated that at least one in seven pregnant women in the USA do not self-report their smoking [81, 82]. Further, a confounding outcome observed in many of the UK-based studies we reviewed, smoking rates in the outcome-affected groups were lower than the unaffected group [9, 55, 83]. Smoking rates reported in UK-based studies tended to be significantly lower than [23] and other USA-based studies that we reviewed, with some UK-based studies reporting no smokers at all [14]. One final issue is that vaping is rapidly replacing smoking, and there has been very little long-term research into the effects of vaping on pregnancy outcomes [84]. For these reasons we chose to leave smoking out of the current model and do not believe its inclusion in this example would significantly alter the result.

**Figure 8:**
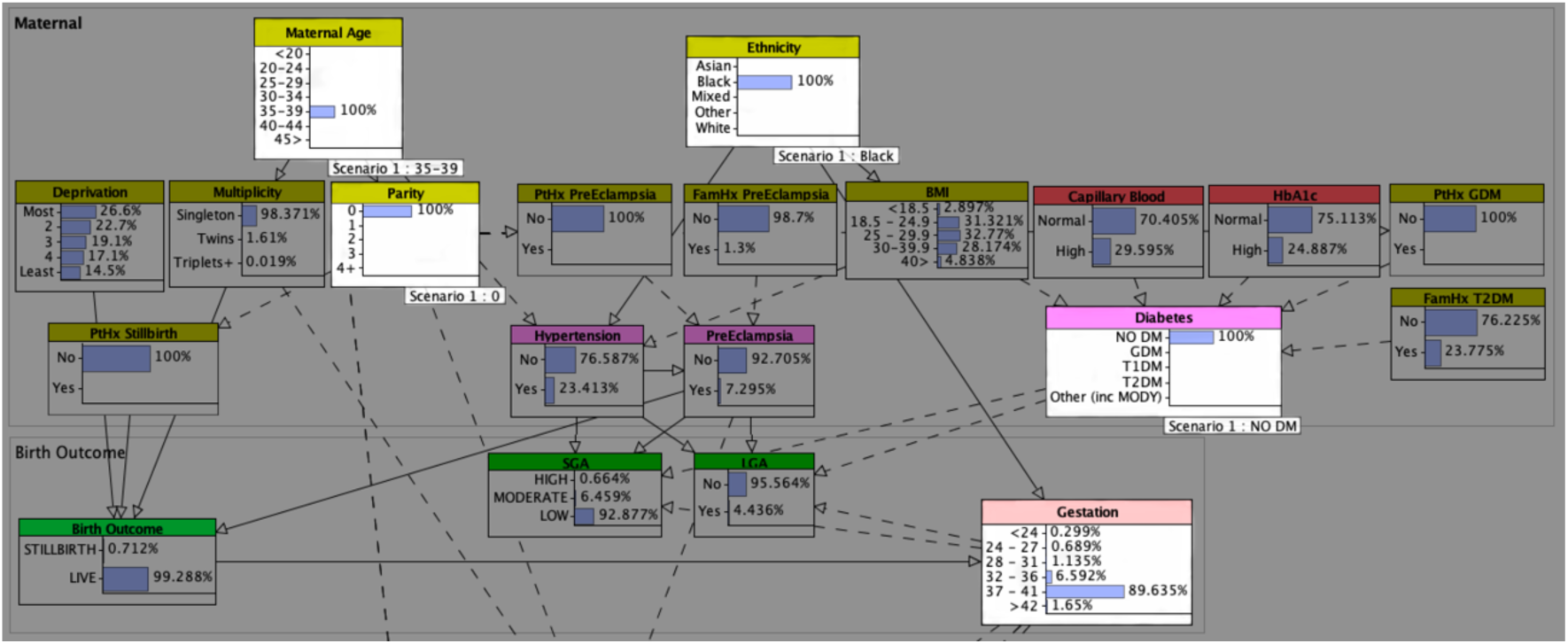
Observation of elements from the vignette of [23]

Finally, [85] developed a nomogram for prediction of the baby’s survivability in pregnancies complicated by GDM using data collected from 626 Chinese mothers receiving outpatient antenatal care between 2016 and 2019. We used their nomogram to evaluate the following scenario:

> *A 35 year old (22 points) Asian woman at 31 weeks or 217 days gestation (10 points) with a BMI of 35 (25 points), first degree family history of T2DM (10 points), history of GDM in a previous pregnancy (4 points) and a mildly high fasting plasma glucose (FPG) of 6.0 mmol (47.5 points). This gave a total of 118.5 points which their nomogram approximated to 82-83% survivability for the baby.*

As shown in Figure 9, our BN model with these same observations predicted a live birth, birth outcome of 84.674%. The dataset of pregnancies used in [85] were temporally the closest to those used to develop our model, and the resulting predictions are not significantly different.

**Figure 9:**
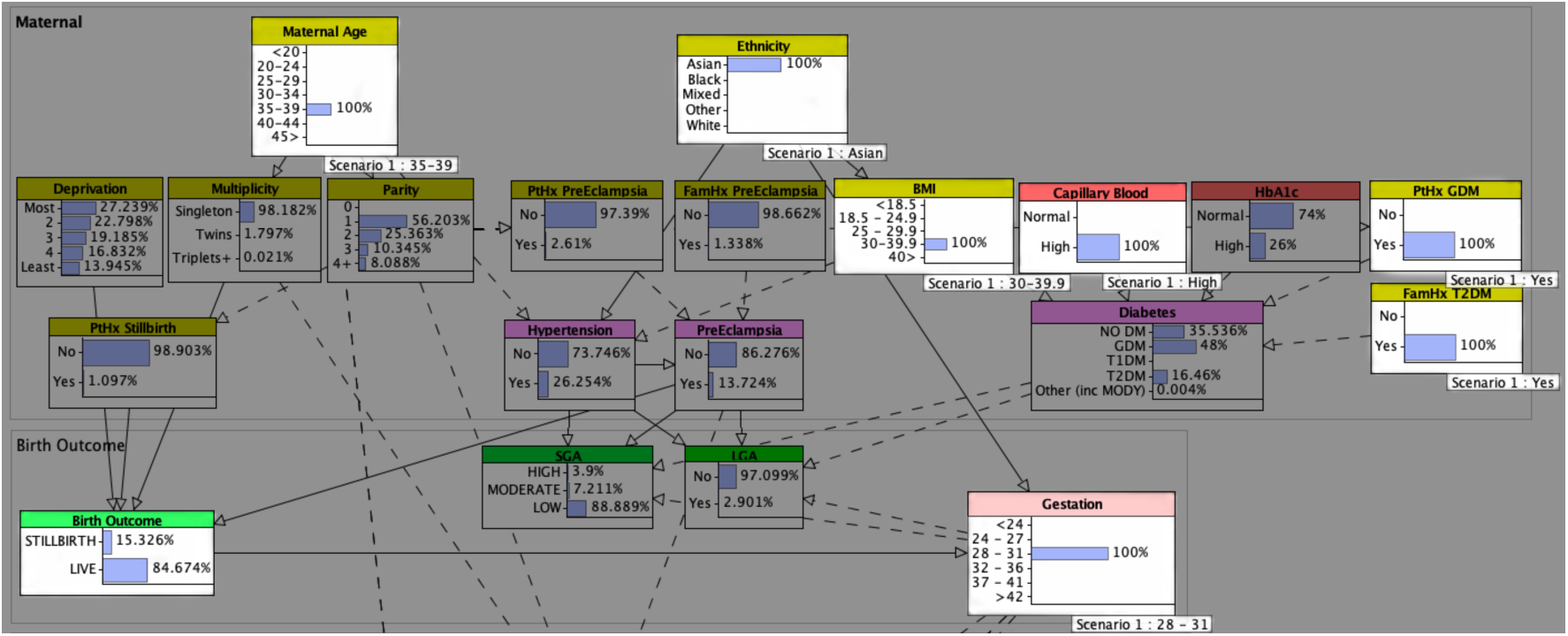
Observation of elements in the [85] comparison

## 5 DISCUSSION

During the process of validating our own model we recognised two further issues arising in the reviewed models not previously discussed in section 2.2. First, was that some of the works we evaluated included elements in their models as risk factors that: (i) are patient self-assessed and based on personal experience of the effector or internalised belief in its effect; (ii) have no absolute method of measurement; or (iii) allow for some degree of manipulation or flexibility when interpreting the result. We refer to these as potentially unquantifiable elements. In the model from [23]*, stress over paying bills* and the woman’s *intent to get pregnant* were examples of patient self-assessed and potentially unquantifiable elements. Second, was the inclusion of elements where the data would be difficult or potentially impossible for the work’s author(s) or patient to reliably procure. We refer to these as *potentially unknowable elements.* In the model from [20] an example of a potentially unknowable element was the mother’s own birthweight when she was born. Families often recorded such things historically, and subsequent generations of a family often grew up and had their children in the same town where they themselves had been born and had access to this knowledge from parents and grandparents. However, this is less common today and what were once common items of information that mothers were told such as the blood group or weight of their baby are less frequently discussed and recorded within the extended family.

While the design process for a model like the one presented in this work may seem resource-intensive and time-consuming, the resulting model is far more efficient in that it does away with the need to enter the same observations for the same patient numerous times into different predictive models. For example, the clinical user might enter *maternal age, parity, maternal weight (or BMI), patient history of gestational hypertension,* and *blood pressure or mean arterial blood pressure* into one model to output a prediction for gestational hypertension [86, 87] - only to later enter many of the same variables including the resulting prediction for gestational hypertension into another model that predicts advancement to preeclampsia [88]. Similarly, the clinical user might enter variables such as *maternal age, maternal weight (or BMI), ethnicity, fasting blood glucose, OGTT results, family and patient history of GDM or DM,* and *current gestational age* into a model that predicts incidence of GDM [89, 90] - only to later enter these and whether or not the mother develops GDM into a model that predicts macrosomia or LGA for the neonate. In both cases we see examples of a one condition that itself is a risk factor for a subsequent condition. The key difference is that when using our model these observations can be entered once for the patient and a probability along the disease pathway for both the primary and subsequent conditions can be computed. Further, when consequent knowledge such as a new risk factor or diagnosis is identified for the patient, these can be observed on the model and the probabilities updated.

Traditionally, clinical lecturers and medical texts describe the normal function of an organ, system or entire organism under observation before progressing to describe how, when and why those natural biological mechanisms may become dysfunctional. Most works that describe development of a model for predicting incidence of a health problem start by describing and modelling either the problem itself, or a mechanism for identifying the problem; yet they rarely demonstrate knowledge or develop a model that describes what it means to be healthy and absent disease. Similarly, most rarely produce a model capable of reasoning from the current observed experience for an entire population and how the disease of interest relates to, or interacts with, other common health issues and known factors. While potentially a valid method for understanding the interaction between the risk factors and symptoms that may lead to one particular disease or adverse outcome, the models produced are only a microcosm of a much larger system, that being the entire patient. The effect of other symptoms or concomitant disease on the disease or outcome of interest cannot be observed. It is not possible with these models to observe the overall effect on the patient when risk factors or symptoms are common to or interact with more than one disease or adverse outcome. Thus, this information is lost. Authors sometimes acknowledge the potential for greater accuracy and health improvement were a model capable of capturing and reasoning the entire scope and scale of risk for those experiencing the modelled disease [11], even as they go on to claim high levels of accuracy for their own disease-limited approach focused on a small list of factors that themselves may turn out to already be commonly accepted general risk factors for poor obstetric outcomes [10, 11]. The difference realised in the model proposed in this work is that a much broader view of the patient is modelled, which incorporates and accounts for the effect of other pregnancy risk factors, symptoms and conditions on both the condition of interest and the patient as a whole organism.

While we are already working on an adaptation of the same model trained with maternity statistics for New Zealand, there are multiple avenues for other future work. The next steps for this work are to incorporate use of the model to develop treatment selection and outcome counterfactuals that allow us to test alternate hypotheses, such as: *what would the outcome be for this patient if we did this? or what would the outcome have been if we hadn’t done that?*

In general, the key *limitations* of the approach used to construct this model may include: (i) availability of sufficiently granular national health statistics for the condition and patient population being modelled, preferably all based on the same population during the same time period; and (ii) access to experts to support model development and face validity assessment. This work was fortunate to have direct access to the clinical expertise throughout the entire project due the fact that the issues and outcomes of maternity had received considerable attention prior to this work such that several large public datasets are regularly published. However, we could foresee these limitations in many other contexts where one would seek to undertake a similar modelling task. Considerable time and resources are required to research, develop, and validate complex and comprehensive models like the one presented here, which may explain why many still prefer to create single-condition, small-factor-count statistical models.

## 6 CONCLUSION

This work has presented a new approach for pregnancy risk prediction modelling that resolves the limiting factors of existing pregnancy outcome modelling approaches. First, rather than developing and training yet another model that is only aware of the condition of interest, our approach delivers a holistic model aware not only of what is but also what is not the condition of interest, including other related conditions and health outcomes. Second, rather than being compromised by the lack of access to sufficient high quality data, our approach is novel in its use of publicly available national health, disease and outcome statistics published by government health departments and care provider networks. The key benefit is that these datasets can be used to develop and train a model both absent of or in conjunction with local retrospective patient cohort data. Third, we used a causal Bayesian probabilistic approach that addresses the issue, and reliably reasons in the presence, of uncertain or missing data. Fourth, we have gone beyond internal statistical validation to use approaches considered more capable of interrogating expert elicited BN models through the use of ongoing face, content and concurrent validation. We found that the model presents an ostensibly accurate description of pregnancies and associated outcomes at both national and individual levels. Finally, and unlike most works, we present three case vignettes with exemplar predictions that can be used by other researchers in comparative validation testing of future models. Our use of concurrency validation also found that as the datasets of pregnancies used in models by authors whose works we used as comparators get temporally closer to the 2021 national maternity datasets our model was learned from, the resulting outcome predictions were also converging.

In conclusion, this work has demonstrated that it is possible to achieve high quality models that are robust and clinically holistic. These are achieved at low cost through the use of causal Bayesian networks together with validation methods that go beyond statistical representations to test the actual structure and reasoning processes and can contribute to instilling a sense of confidence in both the clinical user and patient.

## Data Availability

All data used is publicly available from national/health department sources. All links to these sources are provided in the manuscript

## ACKNOWLEDGMENTS

Acknowledgments are placed before the references. Add information about grants, awards, or other types of funding that you have received to support your research. Author can capture the **grant sponsor information**, by selecting the grant sponsor text and apply style ‘GrantSponsor’. After this, select grant no and apply ‘GrantNumber’ from style panel. Example of Grant sponsor: Competitive Research Programme and example of Grant no: CRP 10-2012-03.

## 7 HISTORY DATES

In case of submissions being prepared for Journals or PACMs, please add history dates after References as (*please note revised date is optional*):

Received November 2019; revised August 2020; accepted December 2020

